# How effective are commercialized products used in the treatment of superficial partial-thickness burn injuries? – Systematic Review with Meta-analysis

**DOI:** 10.1101/2022.11.14.22282317

**Authors:** Luana Nice da Silva Oliveira Merola, Guilherme Albuquerque Sampaio, Marina Clare Vinaud, Erika Lima Coutinho, Ruy de Souza Lino Junior

## Abstract

Burns are an important public health issue, related to 11 million cases annually around the world and up to 180 thousand direct or indirect deaths. Thus, it is vitally important to understand the effectiveness of the different products commercialized for the treatment of burn injuries.

**Objectives:** To analyze the effectiveness of products used in the treatment of superficial-type partial-thickness burn injuries.

**Methods:** This is a systematic review, using the PICO strategy, with a search period between 2004 and 2020, consulting the COCHRANE Library, Lilacs, Medline, PubMed and Scielo databases. The inclusion criteria were studies that used commercialized products for the treatment of superficial-type partial-thickness burn injuries in humans. To assess the quality and risk of bias of the studies, the Oxford scale and criteria from the Cochrane Guidelines were used.

**Results:** 19 eligible studies were selected, most of the products were presented as an alternative to the traditional treatment that involves the use of the silver sulfadiazine product. The methodological quality of the studies allowed performing a meta-analysis of only 2 studies, evaluating the healing outcome, the low number of studies included for statistical analysis suggests that it is not possible to conclude which product is more effective.

**Conclusions:** There is a limitation in the available studies that address the costs and outcomes of existing interventions for the treatment of burns. Future research should develop systematic, valid measures in order to obtain an analytically and statistically adequate result.

## Introduction

Burns are traumatic injuries caused by thermal, chemical, electrical or radioactive agents, causing physiological changes, capable of triggering partial or total destruction of the skin and its annexes, reaching deeper layers, such as subcutaneous tissue, muscles, tendons and bones (Sanches, et al. 2016). Burn injuries are classified by the depth of damage to the anatomical thickness of the skin involved: superficial, partial thickness, full thickness, or subdermal (Abazari, 2022). Superficial burns affect only the epidermis but not the dermis (such as sunburn) and are red, swollen, and painful (Hettiaratchy & Dziewulski, 2004; Andrade, et al. 2010; Abazari, 2022).

Partial thickness burns can be superficial or deep. The superficial ones affect the dermis and its upper layer (papillary dermis) presenting hyperemia and severe pain, leaving minimal scar tissue. Deep partial-thickness burns affect almost the entire thickness of the dermis (reticularis), presenting with a pale, dry color, loss of all epidermal attachments, and less intense pain (Hettiaratchy & Dziewulski, 2004; Inoue, et al. 2016; Abazari, 2022).

In full-thickness burns, the lesion affects the entire thickness of the skin and, in some cases, extends to subcutaneous tissue, muscle, and bone. They present a whitish and rigid appearance and only heal with a graft because there are no dermal elements for regeneration (Andrade, et al. 2010; Yoshino, et al. 2016; Abazari, 2022)

The tissue necrosis resulting from burns provides an environment conducive to microbial growth and subsequent invasion. The severity of this complication is related to the risk factors associated with the patient’s own condition, considering age, the patient’s preexisting diseases, burn depth and wound moisture (Gonella, et al. 2016).

According to the World Health Organization (WHO), approximately 11 million people suffer from burns annually, of which 180,000 die directly or indirectly (WHO, 2018). In develop countries such as USA, 486 thousand health treatments are registered for burns annually (Josephine, 2021). In underdeveloped and developing countries, burns are the fourth most common type of trauma and one of the main causes of mortality and disability (Rigon, et al. 2019).

In Brazil, burns are an important public health problem, as it is estimated that 1 million Brazilians suffer from burn accidents annually, of which approximately 120,000 people generate expenses for the public health service both due to the need for procedures and for post-operative follow-up hospitalization, since psychological and physical sequelae can occur, causing a high rate of disability in addition to the death rate of 2 to 3% directly or indirectly (Morais, et al. 2017; Bezerra, et al. 2020). Regarding the pediatric population, the risk of death from burn wounds is high, with a global rate of 3.9 deaths per 100,000 according to the WHO. In addition to the high number requiring hospitalizations, in Brazil alone in 2021 there were more than 9000 hospitalizations of children and teenagers’ victims of burns, with the main age group affected being 1 to 4 years old (DATASUS, 2021; WHO, 2018).

If the burn wound is not treated properly there is a great risk of developing various infectious complications. In addition to local changes, burn wounds can also lead to systemic disorders, which are caused by pain, loss of blood plasma, tissue proteins breakdown by the body and hypermetabolism. The current challenge is choosing the ideal treatment method to accelerate healing and attenuate complications (Markiewicz-Gospodarek, et al.2022; Stanojcic, et al. 2022).

It is of vital importance to understand the different commercially available products to combat infectious processes arising from traumatic injuries, since the non-intact skin is the main gateway for microorganism’s invasion. Secondary infections are considered the main causes of death of the severely burned patient (Lau, et al. 2016).

The healing process is essential for the survival of the organism and may occur through regeneration or healing. The healing process fundamentally occurs in three phases: 1. inflammation, 2. formation of granulation tissue and 3. maturation and remodeling of the extracellular matrix (Oliveira and Dias, 2013).

The most striking feature of the final phase of wound healing is the large and accelerated synthesis and degradation of collagen in the wound region for the formation of new extracellular matrix. Thus, there is the presence at the site of metalloproteinase enzymes (MMPs), metalloproteinase inhibitors (TIMPs) produced by various inflammatory, endothelial, fibroblast and keratinocyte cells. The balance between the productive action of these cells is important to prevent the formation of hypertrophic scars and keloids (Takeo, et al. 2015).

There are several ways to assess wound healing. Currently, the most used methods are densitometry, immunohistochemistry, microscopic and macroscopic analyses, collagen morphometry, tensiometry, and, most recently, the dosage of growth factors (Campos et al. 2007; Ferreira and Paula, 2013; Babakhani, et al. 2020).

The commercialized product available for health institutions most used in the treatment of burn wounds is based on silver sulfadiazine. It is an efficient topical antimicrobial agent. However, other products that stimulate healing should also have its use stimulated, such as Aquacel®, Mepilex® among many others. The knowledge of the existence, efficacy and mode of action of alternative therapies improve the proper management of burn wounds in clinical practice (Ferreira & Paula, 2013).

There are several new therapies that may offer potential benefits to the healing process, however there is a limited number of preclinical studies concerning their applicability and results, many of them with methodological limitations. This limited number of studies can bring uncertainty in decision-making for health managers and professionals regarding the incorporation of these new technologies (Mizuno, 2010; Oliveira, 2011).

The study of the effectiveness of commercialized products will establish parameters for health managers in decision-making for health care in the treatment of burns through systematic protocols, it will also help the scientific community to observe the need for more research in the area for the theoretical basis (Mizuno, 2010).

Systematic Review and Meta-analysis are scientific methodological tools based on evidence. They are secondary studies in which there is a defined methodology, the objective is to synthesize and evaluate the results of primary studies simultaneously (Brasil, 2014). The aim of this study is to propose a systematic review with meta-analysis to offer scientific subsidies regarding the adequate products used in burn wounds care.

## Methodology

A systematic review was carried out with a meta-analysis regarding the efficacy and effectiveness of products used in the treatment of superficial-type partial-thickness burn injuries. It was developed by 2 researchers from the IPTSP (Institute of Tropical Pathology of Tropical Health) at the Federal University of Goiás (UFG).

The systematic review (SR) was registered in PROSPERO on May 28, 2020, n. 211777. A protocol was developed in order to formalize, monitor and document all the steps to be carried out in SR (Berwanger, 2007).

It was necessary to use the PICO technique, for the construction of the question, this technique is an acronym for Patient, Intervention, Comparison and “Outcomes”. The guiding question of the systematic review was: What is the effectiveness of the products used in the treatment of partial-thickness burn injuries? (Systematic Review) Applying the PICO strategy:

P: population/patients – patients who present burn injuries.

I: event / intervention – commercialized products used in the treatment of such wounds

C: comparison/ control – there are no control, only the products described in the selected articles, according to each study and methodology.

O: outcome – analysis of te efficacy on the healing of the burn wound.

### Article search and selection strategy

The search for articles in the literature was performed in the following databases: Medline (via Pubmed), Virtual Health Library (BVS), The Cochrane Library Center for Reviews and Dissemination (CRD), Lilacs and Scielo.

The advanced search mode was used in order to insert the descriptors (DeCS) (burns AND products burns AND treatments), in addition to the selection of “all fields”.

There was a search in the gray literature through the search in the references of the cited articles, to find more articles related to the theme.

Studies in English, Spanish and Portuguese were selected, studies were searched from January 2004 to December 2020 was carried out, according to the SR guideline.

The inclusion criteria were studies that used commercialized products for the treatment of superficial-type partial-thickness burn injuries in humans.

A free reference manager, Mendeley, was used for sorting articles, accounting for duplicates, organizing references, practicality and time optimization.

After an initial triage, the studies that were considered eligible were read in full. Eligible studies were those that showed the efficacy of products used for partial burns only in humans, with no criteria for selection of sex, race or age.

Articles selection and screening were performed independently by two researchers and the results were compared. Disagreements were resolved in consensus meetings or by arbitration through a third researcher.

### Data extraction

The two researchers independently assessed the compliance of full texts with the inclusion criteria, identified the names of authors, institutions, year of publication, scientific journals, outcome when applying the eligibility criteria.

A data extraction form was prepared, in accordance with the guidelines for analyzing the evidence of marketed products found in the studies, and they were presented in a table in the results section, in order to allow comparison of the parameters selected for efficacy (Supporting Information 1). The Oxford Scale (Nobre, 2006) was applied by two authors independently, to assess the quality of the evidence of the articles, in this criterion, the types of studies are evaluated: articles published in indexed journals with evidence level 1A, 1B, 1C, 2A, 2B, 2C, 3A, 3B and 4, according to the classification of the Oxford Center for Evidence-Based Medicine (Sampaio, 2007).

### Study variables/analyzed variables

The outcome was to analyze the effectiveness by the period of time in order for the injury to close and the burn healing rate by commercialized products. Through the Pressure Ulcer Scale for Healing (PUSH) tool in which the primary outcome is the appearance of the wound, resulting in complete healing and in its minimum time of weeks and the secondary outcomes were to analyze the reduction in percentage of the wound area and percentage of change in the area in the minimum time of weeks (Garbuio, 2018).

Quantitative variables were also evaluated in studies that can be applied to meta-analysis in binary outcomes, choosing as a control the comparison to the drug silver sulfazianide.

### Statistical analysis

Data meta-analysis was performed using the R software version 4.2.1. Within it, the Meta package was chosen to run the data and the metabim command to perform a meta-analysis of categorical outcomes, using the random effect model, presented as Relative Risk (RR), with a confidence interval of 95% (CI) and statistical significance when p<0.05.

Heterogeneity was evaluated using the I² statistic, where an I² value close to 0% indicates no heterogeneity between studies, close to 25% indicates low heterogeneity, close to 50% indicates moderate heterogeneity and close to 75% indicates high heterogeneity between studies.

The analyzes classified as Relative Risk, the following methods were used: *Mantel-Haenszel method and Restricted maximum-likelihood estimator for tau^2*. Results were displayed in Forest Plot chart.

## Results

674 references were identified, 648 in the data sources and 26 through manual searches of the references. After the exclusion of duplicate publications and selection by title and abstract, at the end, 19 articles met the inclusion criteria and were analyzed in full, as shown in Supporting Information 2 and Figure 1.

**Figure 1.**
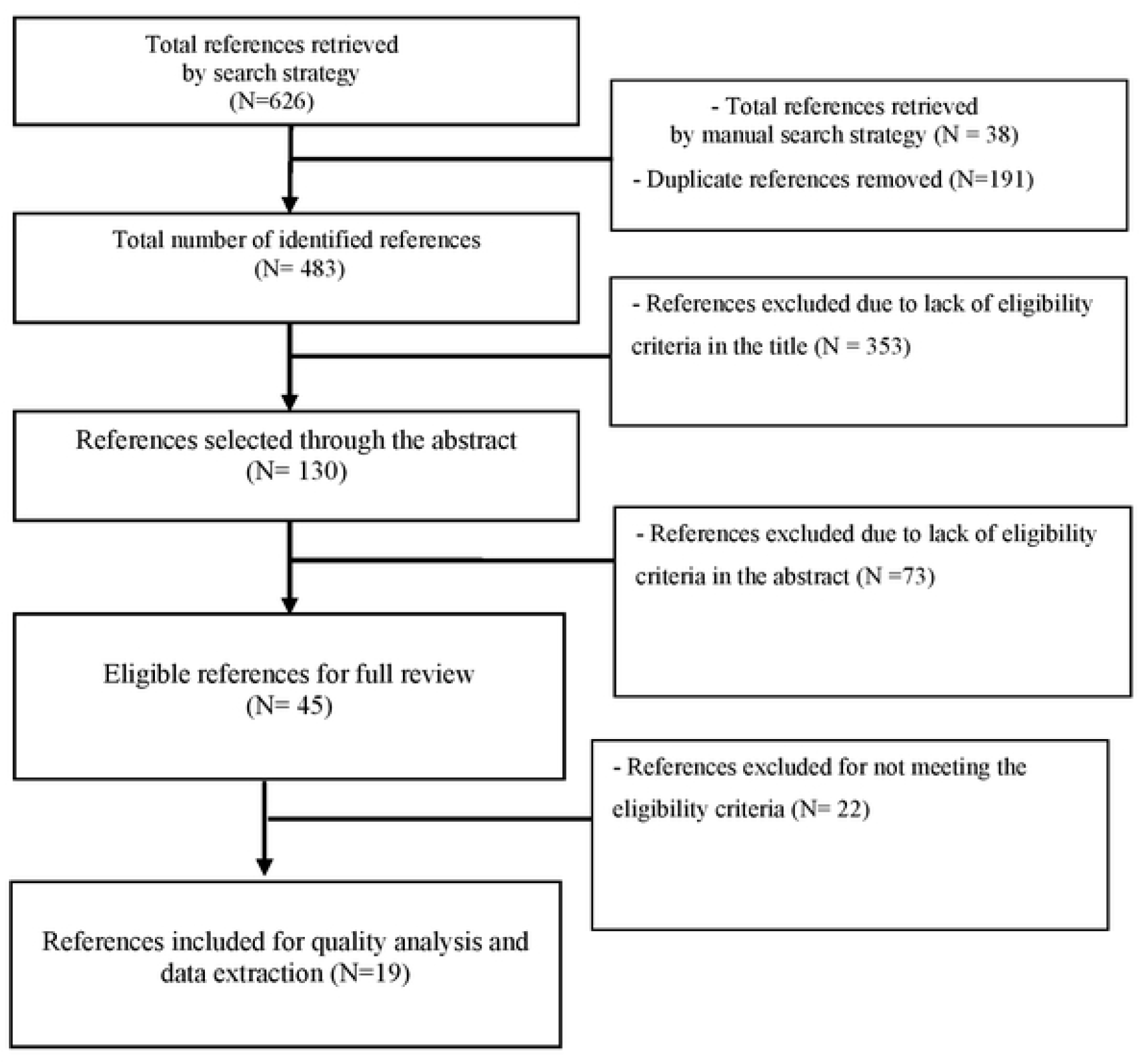
Flowchart of the article selection process

Of these articles, five studies were classified as a cross-sectional study, classified as 2B in level of evidence, and two were characterized as a systematic review, classified as level 2A according to the criteria of the Oxford Center for Evidence-Based Medicine classification.

In this study, it was observed that most of the articles are recent, published mainly from 2011, the last two being from 2020, which can be seen in Supporting Information 2, as well as showing an overview of all studies included in the final sample and of all data elements used in the data analysis process.

Among the 19 included studies, 2 evaluated the outcome of the use of other products for burns (keratin and hydrogel; in relation to silver sulfadiazine, relating wound healing time). The meta-analysis demonstrated by the forest plot graph (Figure 2) showed that the use of alternative products in the literature showed a reduction of about 33% (1-RR) in wound healing time compared to the use of silver sulfadiazine (RR 0.67 / 95% CI 0.48-0, 93 / p<0.01). The I² statistic indicated no heterogeneity between the studies (I²= 0%)

**Figure 2.**
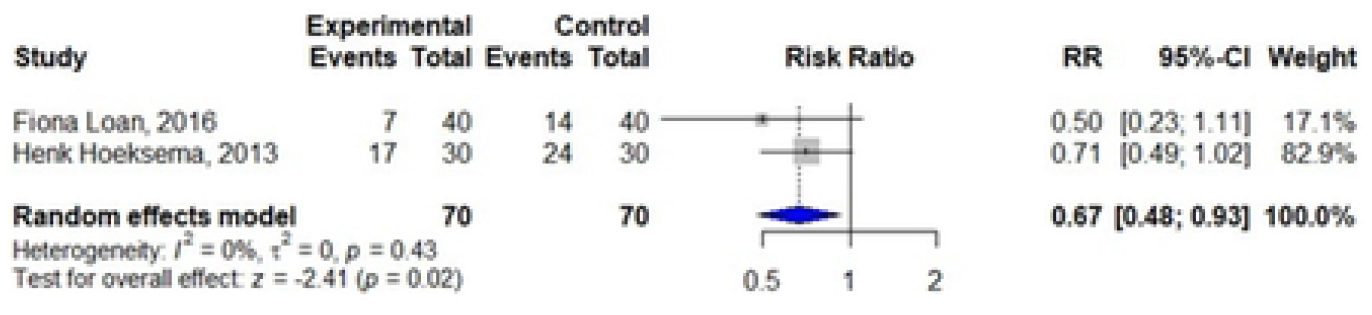
Forest Plot of studies included in the systematic review that compare their productS wilh the use of silver sulfadiazine and their respective association measures

## Discussion

Of the 19 articles selected in this review, 7 articles describe randomized clinical trials. It is worth mentioning the presence of two systematic reviews, one of them also involving meta-analysis. The fact that only 19 articles were selected for this review, despite the search covering a period of a decade, demonstrates a scarcity of studies related to the treatment of burns.

In these studies, we sought to assess, for different interventions, the length of hospitalization and dressing changes, the levels of pain and discomfort, bacterial load of the wound, healing and the time required for total re-epithelialization. The satisfaction of health professionals and guardians, in the case of patients who needed care such as children, regarding the handling of the dressing and the economic cost during treatment was also taken into account.

Most interventions were presented as an alternative to traditional treatments involving daily change dressings using silver sulfadiazine or other antibiotics. Most new interventions seek to reduce wound handling by reducing dressing changes, leading to lower levels of patient pain and discomfort and faster re-epithelialization. The adverse effects of traditional silver-based drugs, such as ointments containing 1% silver sulfadiazine, were also motivating for investigation of alternative ways of transporting this metal (HOEKSEMA, et al. 2013; NHERERA, et al. 2017).

Among the interventions presented, the Biobrane® dressing stood out. This dressing was investigated for its effectiveness in seven articles, and the treatment was recommended by four of them. In a systematic review, whose objective was to select the best treatment for children with burns considering re-epithelialization and scar formation, among the analyzed treatments, Biobrane® showed a better epithelialization rate, reduced hospitalization period and reduced pain (VLOEMANS, et al. 2014).

Feng et al. (2018) reported the cases of 2 patients with superficial partial-thickness burns in the pubic region. Healing time without morbidity and pain rate when using Biobrane® dressing and daily dressings were compared. After 7 days, it was reported that the use of the Biobrane® dressing reduced the regular dressing changes, eliminating the pain caused by such handling. This same dressing was investigated by Whitaker et al. (2007) in a report of a series of cases of 32 burn units in the UK between October and November 2005. Patients were followed for a minimum of 48 hours. The article deals with the use and recommendations of Biobrane® in burn units in the UK. It was reported that, during the period in which the research was carried out, most burn care units used Biobrane® in the treatment of partial thickness burns in patients of all ages. Silicone is also found in Biobrane® dressing, which is composed of a thin, semi-permeable membrane (there are pores for exudate drainage) attached to a nylon layer (WHITAKER, et al. 2007). This nylon mesh is covered by a layer of type 1 collagen of porcine origin (GREENWOOD, 2011). AWBAT-S® dressing is very similar to Biobrane® in terms of structure. The difference lies in the greater porosity of AWBAT-S® and the absence of potentially harmful compounds used in the covalent bonding of collagen to the Biobrane® dressing (GREENWOOD, 2011). Common dressings (found in hospital pharmacies) containing a collagen membrane were also employed in some of the studies (RAMAKRISHNAN, et al. 2013; SINGH, 2020).

The efficacy of Biobrane® was also tested against the use of AWBAT-S® in a prospective randomized study carried out by Greenwood in 2011. In it, patients presented superficial burns to partial superficial burns with spontaneous healing, with 2 to 40% of total body surface area (TBSA). They were treated with one of the approaches and followed up for the next 12 months. The following were evaluated: length of hospitalization period, patient-reported pain perception, time to heal, scarring and cost of care for treated patients. AWBAT-S® proved to be better than Biobrane® in terms of ease of removal and discomfort experienced by patients at this time, and is at least as good in terms of length of hospitalization period, total healing time, and pain/discomfort experienced by patients during rest healing and therapy.

In 2013, Ramakrishnan et al. published a clinical trial that demonstrated the advantages of collagen membranes in wound healing. This trial, conducted from 1992 to 2011, included 487 pediatric patients with superficial and deep partial-thickness burns and mixed-thickness burns at Kanchi Kamakoti Childs Trust Hospital. The different types of collagen dressings used proved to be highly advantageous for burn patients in developing countries as they adapt to humid and hot environments and are cost-effective. This was the study with the largest number of patients among all the articles analyzed in the present work.

Seven years later, Singh *et al*. (2020) published a randomized clinical trial that compared the outcome of treatment with collagen dressings and paraffin gauze associated with silver sulfadiazine. 68 patients with fresh acute superficial partial burns were divided between the two groups and followed up for clinical evolution until the burns healed. 90% of patients in a collagen dressing group did not need any or only one dressing change, with a shorter hospital stay. In paraffin gauze plus silver sulfadiazine group, 22 cases required 3 to 5 dressing changes.

Many dressings have silver embedded in their structure, either in the form of nanocrystals or in its ionic form. The Aquacel® Ag dressing has 1.2% of ionic silver in a hydrofiber base composed of sodium carboxymethylcellulose, in the form of a hydrated gel that activates the silver ions to exert an antimicrobial effect. It was addressed in three articles and recommended in two of these and was the second most cited intervention in the articles used in this study (YARBORO, 2013; LAU, et al. 2016; AGGARWALA, et al. 2020;).

In the study published by (Yarboro *et al*. in 2013, a randomized clinical trial was performed, which included 24 patients (ages 19 to 53) with superficial partial-thickness burns of 0 to 4 days old and less than 25% TBSA. The study established a comparison between the Aquacel® Ag dressing and the traditional silver sulfadiazine dressing. Patients were followed until complete healing. Both dressings had similar results in terms of effectiveness, however pain was reduced with Aquacel® Ag, as well as the number of treatments received in order to achieve a 100% re-epithelialization rate.

Lau et al. published, in 2016, a retrospective review of the treatment of 119 patients admitted with burns between January 2010 and December 2014, with TBSA of 2 to 25%. These were treated with either Aquacel® Ag or the paraffin gauze dressing and followed up for 5 years to assess the effectiveness of Aquacel® Ag. This appeared to promote early healing of burn wounds with less hypertrophic scar formation, shorter hospitalization period, and much less frequent dressing changes compared to paraffin gauze dressings.

Furthermore, Aquacel® Ag, along with Acticoat® and Biobrane®, was compared to Mepilex® Ag in a randomized clinical trial performed by Aggarwala et al. (2020). In it, 131 partial-thickness burn wounds in 119 patients were randomized and those treated with Mepilex® Ag achieved faster re-epithelialization with better cost-effectiveness.

Another intervention consisted of gels for topical use, based on keratin, with diversity in thicker consistency (Keragel®), intermediate (Kematrix®) and thinner (KeragelT®). The effectiveness of these products were also compared to treatment with Biobrane® together with Acticoat® and keratin products were found to be well tolerated with minimal pain, itching and scarring, as well as being cost effective and easy to use by the healthcare professional in a cohort study of 80 partial thickness burn patients within 24 hours (Loan et al. 2016).

Another gel used as an alternative had 10% birch bark triterpene dry extract (about 72 to 88% betulin) (FREW, et al. 2019), marketed under the name Oleogel-S10® presented in an assay randomized clinical trial to accelerate the healing process, in addition to improving skin texture, pigmentation and redness compared to the standard wound intervention, Octenilin® gel, which contains silver. A 12-month follow-up involving 61 patients with superficial burns of partial thickness caused by fire, heat or scalding within 48 hours of injury. Suprathel®, an alloplastic copolymer of polyactide, trimethylene carbonate, and ε-caprolactone, developed as a synthetic skin substitute, was another dressing cited to treat superficial partial-thickness burns in pediatric patients (HIGHTON, et al. 2013; EVERETT, et al. 2015;).

Cellulose sheets of microbial origin and a dressing composed of a polyester weft impregnated with a matrix of hydrocolloid particles (carboxymethylcellulose), petrolatum and polymers for cohesion, known as Urgotul®, were also used que demonstraram serem mais eficazes do que than a silver sulphadiazine cream for treatment of partial thickness burns (LETOUZE, et al. 2004; ABOELNAGA, et al. 2018).

The use of a topical ointment containing a protein complex was also described, formed by a glucose oxidase and a lactoperoxidase, stabilized in polymers contained in a matrix of water and polyethylene glycol, marketed as Flaminal® (HOEKSEMA, et al. 2013). Treatment with Flaminal® showed shorter hospitalization period and better healing quality through a retrospective cohort study that compared length of hospitalization period, bacterial load and time to wound closure when the burn was treated with Flaminal® and when Flammazine® (1% silver sulfadiazine) was used. In total, 60 hospitalized outpatients with burns were treated with Flaminal® or Flammazine® between 1998 and 2003, excluding superficial burns and burns treated with skin grafts.

Finally, there was an intervention with the use of *Aloe vera* gel for the treatment of burns, which showed an advantage in terms of early wound re-epithelialization, early pain relief and a better cost-benefit compared to 1% silver sulfadiazine cream (SHAHZAD; AHMED, 2013).

As for costs, only four articles cited the necessary expenditure for each intervention. The total cost of treatment with keratin-based drugs was US$2635.00 per patient, compared to US$5016.00 spent with the traditional approach (LOAN, et al. 2016). Aggarwala et al. (2020) reported an average cost of USD$115.92 for treatment with Biobrane®, USD$146.39 when Acticoat® was used, USD$36.62 for treatment with Aquacel® Ag, and USD$60.00 for treatment with recommended treatment with Mepilex® Ag.

Among the periods required for complete post-treatment re-epithelialization, the lowest average was 7.6 days for wound closure with the use of Oleogel-S10® (FREW, et al. 2019). The longest time required for healing was with the use of Suprathel®, with an average of 24 days for complete re-epithelialization (RAHMANIAN-SCHWARZ, et al. 2011). Both studies reported burns that are similar in depth and thickness, but include discrepant patient numbers (61 and 34 respectively), although both are quite limited. This small number of patients is a characteristic observed in most of the articles described here, evidencing the lack of large studies such as the one reported by Ramakrishnan et al. (2013).

Another study with Aquacel® Ag had a cost of USD$7.00 per 0.01 m2 of dressing used, being more expensive than traditional dressings, but saving USD$4000.00 in expenses related to frequent dressing changes and hospitalization (LAU; WONG; TAM, 2016). Treatment with *Aloe vera* gel was also priced, with 1000 mL of the gel being purchased for USD$6.20 compared to USD$7.96 for each 250 g of silver sulfadiazine cream (SHAHZAD, et al. 2013). The cost of dressing per percentage of burned surface was USD$0.064 for 2 g of cream and USD$0.03 for 5 mL of gel. The other articles did not mention concrete amounts of expenditure on interventions.

## Conclusion

The scarcity of large studies that address the costs and results of existing interventions for the treatment of burns makes it difficult to carry out a survey of relevant data on the best treatments available on the market.

In addition, two studies were funded by the laboratory that manufactures the drug used (studies involving Oleogel-S10® and Urgotul®) and another had the dressings donated by the factory (microbial cellulose sheets), increasing the chances of bias in the results presented and placing the reliability of the studies in check (LETOUZE, et al. 2004; ABOELNAGA, et al. 2018; FREW, et al. 2019).

As for the meta-analysis analysis, it is suggested that the low number of studies included for statistical analysis, because of the heterogeneity in the methodologies of the evaluated studies, it is not possible to conclude that other alternatives are actually more effective than silver sulfadiazine.

## Data Availability

All relevant data are within the manuscript and its Supporting Information files.

## References

Abazari M, et al. A systematic review on classification, identification, and healing process of burn wound healing. Int. J. Low. Extrem. Wounds. 2022 doi: 10.1177/1534734620924857.

Aboelnaga A, et al. Microbial cellulose dressing compared with silver sulphadiazine for the treatment of partial thickness burns: A prospective, randomised, clinical trial. Burns, v. 44, n. 8, p. 1982–1988, dez. 2018. doi.org/10.1016/j.burns.2018.06.007

Andrade AG, Lima CF, Albuquerque AKB. Efeitos do laser terapêutico no processo de cicatrização das queimaduras: uma revisão bibliográfica. Rev Bras Queimaduras, v.9 2130, 2010. Disponível em: http://rbqueimaduras.com.br/details/29/pt-BR/efeitos-do-laser-terapeutico-no-processo-de-cicatrizacao-das-queimaduras--uma-revisao-bibliografica

Atiyeh BS, Hayek SN, Gunn SW. New technologies for burn wound closure and healing: review of the literature. Burns. 2005;31(8):944–56. doi: 10.1016/j.burns.2005.08.023

Babakhani A, et al. Effects of Hair Follicle Stem Cells on Partial-Thickness Burn Wound Healing and Tensile Strength. Iran Biomed J. 2020 Mar;24(2):99–109. doi: 10.29252/ibj.24.2.99. doi: 10.29252/ibj.24.2.99

Berwanger O, et al. Como avaliar criticamente revisões sistemáticas e metanálises?. Rev. bras. ter. intensiva [Internet]. 2007; 19(4):475–480. Disponível em: http://www.scielo.br/scielo.php?script=sci_arttext&pid=S0103-507X2007000400012&lng=en.

Bezerra AFC, et al. Mortality to burns in children between zero and four years in Brazil. Braz. J. of Develop, 6 (7), 43012–43023. 2020. https://doi.org/10.34117/bjdv6n7-062

Brasil. Ministério da Saúde. Secretaria de Ciência, Tecnologia e Insumos Estratégicos Departamento de Ciência e Tecnologia. Diretrizes Metodologicas, Elaboração de revisão sistemática e metanalise de estudo observacionais comparativos sobre fatores de riscos e prognósticos – Brasília : Ministério da Saúde, 2014; 33p.

Campos ACL, Borges-Branco A, Groth AK. Cicatrização de feridas. Arq Bras Cir Dig 2007; 20:51–58. doi.org/10.1590/S0102-67202007000100010

Evertt M, et al. Use of a copolymer dressing on superficial and partial-thickness burns in a paediatric population. Journal of Wound Care, v. 24, n. Sup7, p. S4–S8, jul. 2015. doi: 10.12968/jowc.2015.24.Sup7.S4

Evers LH, Bhavsar D, Mailänder P. The biology of burn injury. Exp Dermatol 2010; 19: 777783. doi: 10.1111/j.1600-0625.2010.01105.x

Feng JJ, See J, Choke A, et al. Biobrane™ for burns of the pubic region: minimizing dressing changes. Military Med Res 5, 29 (2018). doi/:org/10.1186/s40779-018-0177-2

Ferreira FV, Paula LB. Sulfadiazina de prata versus medicamentos fitoterápicos: estudo comparativo dos efeitos no tratamento de queimaduras. Rev Bras Queimaduras 2013; 12: 132139. Disponível em: http://rbqueimaduras.org.br/details/158/pt-BR/sulfadiazina-de-prata-versus-medicamentos-fitoterapicos--estudo-comparativo-dos-efeitos-no-tratamento-de-queimaduras

Frew Q, et al. Betulin wound gel accelerated healing of superficial partial thickness burns: Results of a randomized, intra-individually controlled, phase III trial with 12-months follow-up. Burns, v. 45, n. 4, p. 876–890, jun. 2019. doi: 10.1016/j.burns.2018.10.019

Garbuio DC, et al. Instrumentos para avaliação da cicatrização de lesões de pele: revisão integrativa. Rev. Eletr. Enf. [Internet]. 20: v20a40. 2018. doi: http://dx.doi.org/10.33448/rsd-v10i11.19246

Galvão CM, Sawada NO, Trevizan MA. Revisão sistemática: recurso que proporciona a incorporação das evidências na prática da enfermagem. Rev Latino-am Enfermagem ; 12(3):549–56, 2004. doi.org/10.1590/S0104-11692004000300014

Gonella HA., et al. Análise da microbiota bacteriana colonizadora de lesões provocadas por queimaduras nas primeiras 24 horas. Revista da Faculdade de Ciências Médicas de Sorocaba, [S.l.], v. 18, n. 1, p. 19–23, xabr. 2016. ISSN 1984-4840. doi.org/10.5327/Z1984-4840201621885

Greenwood JEA, Randomized, Prospective Study of the Treatment of Superficial Partial-Thickness Burns: AWBAT-S Versus Biobrane. Eplasty, v. 11, p. e10, 2011. Disponível em: https://www.ncbi.nlm.nih.gov/pmc/articles/PMC3045219

Josephine A. and Shahriar S. Burn Infection and Burn Sepsis. Surgical Infections. 22:1, 58-64. 202. doi: 10.1089/sur.2020.102

Highton, et al. Use of Suprathel® for partial thickness burns in children. Burns, v. 39, n. 1, p. 136–141, fev. 2013. doi.org/10.1016/j.burns.2012.05.005

Hoeksema H, et al. A comparative study of 1% silver sulphadiazine (Flammazine®) versus an enzyme alginogel (Flaminal®) in the treatment of partial thickness burns. Burns, v. 39, n. 6, p. 1234–1241, set. 2013. doi: 10.1016/j.burns.2012.12.019

Inoue Y, et al.; Wound/Burn Guidelines Committee. The wound/burn guidelines - 1: Wounds in general. J Dermatol. 2016 Apr;43(4):357–75. doi: 10.1111/1346-8138.13276. Epub 2016 Mar 12. PMID: 26972819.

Isaac C, Ladeira PRS, Rêgo FMP, Aldunate JCB, Ferreira MC. Processo de cura de feridas: cicatrização fisiológica. Rev Med (São Paulo) 2010; 89: 125–131. doi.org/10.11606/issn.1679-9836.v89i3/4p125-131

Lau, C. T. e Wong, k. K.Y. e Tam, P. Silver containing hydrofiber dressing promotes wound healing in paediatric patients with partial thickness burns. Pediatric Surgery International. 32: 577–581, 2016. Doi: 10.1007/s00383-016-3895-0

Letouze A, et al. Using a new lipidocolloid dressing in paediatric wounds: results of French and German clinical studies. Journal of Wound Care, v. 13, n. 6, p. 221–225, jun. 2004. doi: 10.12968/jowc.2004.13.6.26630

Loan F, et al. Keratin-based products for effective wound care management in superficial and partial thickness burns injuries. Burns, v. 42, n. 3, p. 541–547, maio 2016. doi: 10.1016/j.burns.2015.10.024

Markiewicz-Gospodarek A, et al. Burn Wound Healing: Clinical Complications, Medical Care, Treatment, and Dressing Types: The Current State of Knowledge for Clinical Practice. Int J Environ Res Public Health. Jan 25;19(3):1338. 2022. doi: 10.3390/ijerph19031338

Mathangi RK, et al. Advantages of collagen based biological dressings in the management of superficial and superficial partial thickness burns in children. Annals of Burns and Fire Disasters, v. 26, n. 2, p. 98–104, 2013. Disponível em https://pubmed.ncbi.nlm.nih.gov/24133405/

Mitsunaga JJK, et al. Rat an experimental model for burns: A systematic review. Acta Cir. Bras. 2012. Disponível em: http://www.scielo.br/scielo.php?script=sci_arttext&pid=S0102-86502012000600010&lng=en.

Mizuno CH. O custo da obesidade no Brasil: A importância da avaliação econômica na tomada de decisão em políticas públicas de prevenção em saúde. 2010; Disponível em: http://www.each.usp.br/flamori/images/TCC_Cintia_2010.pdf

Morais IH, Daga H, Prestes M. A. Crianças queimadas atendidas no Hospital Universitário Evangélico de Curitiba: perfil epidemiológico. Rev Bras Queimaduras, 15 (4), 256–260. 2017. Disponível em: http://www.rbqueimaduras.com.br/details/323/pt-BR/criancas-queimadas-atendidas-no-hospital-universitario-evangelico-de-curitiba--perfil-epidemiologico

Nherera LM, et al. A systematic review and meta-analysis of clinical outcomes associated with nanocrystalline silver use compared to alternative silver delivery systems in the management of superficial and deep partial thickness burns. Burns, v. 43, n. 5, p. 939–948, ago. 2017. doi: 10.1016/j.burns.2017.01.004

Nobre M, Bernardo W. Prática clínica baseada em evidência. Rio de Janeiro: Elsevier; 2006. doi.org/10.1590/S0104-42302004000100045

Oliveira, I; Dias, V. Cicatrização de feridas: fases e fatores de influência. Acta VeterinariaBrasilica, v. 6, n. 4, p.267–271, 2013. Disponível em: https://periodicos.ufersa.edu.br/index.php/acta/article/view/2959.

Oliveira FL, Serra MCVF. Infecções em queimaduras: revisão. Rev Bras Queimaduras; 10(3):96–99, 2011. Disponível em: http://www.rbqueimaduras.com.br/details/71/pt-BR/infeccoes-em-queimaduras--revisao#:~:text=Organismos%20associados%20a%20infec%C3%A7oes%20em,na%20primeira%20semana%20de%20interna%C3%A7ao.

Rahmanian-Schwarz A, et al. A clinical evaluation of Biobrane® and Suprathel® in acute burns and reconstructive surgery. Burns, v. 37, n. 8, p. 1343–1348, dez. 2011. doi: 10.1016/j.burns.2011.07.010.

Rigon AP, et al. Perfil epidemiológico das crianças vítimas de queimaduras em um hospital infantil da Serra Catarinense. Rev Bras Queimaduras, 18 (2), 107–112. 2019. Disponível em: https://pesquisa.bvsalud.org/portal/resource/pt/biblio-1119561.

Sampaio RF, Mancini MC. Estudos de revisão sistemática: um guia para síntese criteriosa daevidência científica. Rev. bras. Fisioter 2007; 11(1): 83–89. doi.org/10.1590/S1413-35552007000100013

Santos CMC, Pimenta CAM, Nobre MRC. A estratégia PICO para a construção da pergunta de pesquisa e busca de evidências. Rev. Latino-Am. Enfermagem 2007. doi.org/10.1590/S0104-1169200700030002.

Santos EJF Dos, Cunha M. Interpretação crítica dos resultados estatísticos de uma meta-análise: estratégias metodológicas. Millenium [Internet]. 2013;44:85–98. Disponível em: http://repositorio.ipv.pt/handle/10400.19/2273.

Shahzad MN, Ahmed N. Effectiveness of Aloe Vera gel compared with 1% silver sulphadiazine cream as burn wound dressing in second degree burns. JPMA. The Journal of the Pakistan Medical Association, v. 63, n. 2, p. 225–30, fev. 2013.Disponível em: https://jpma.org.pk/PdfDownload/4001

Shivani A, et al. Treatment of partial thickness burns: a prospective, randomised controlled trial comparing BiobraneTM, ActicoatTM, Mepilex® Ag and Aquacel® Ag. Journal of Burn Care &Research, p. 1–10, 2020. doi: 10.1093/jbcr/iraa158

Singh A, Bhatnagar A. Management of superficial partial thickness burn with collagen sheet dressing compared with Paraffin Gauze and silver sulfadiazine. Annals of Burns and Fire Disasters, v. 33, n. 3, p. 233–238, 2020. Disponível em: https://www.ncbi.nlm.nih.gov/pmc/articles/PMC7680202/pdf/Ann-Burns-and-Fire-Disasters-33-233.pdf

Stanojcic M, et al. Pathophysiological Response to Burn Injury in Adults. Ann Surg. 2018 Mar;267(3):576–584. doi: 10.1097/SLA.0000000000002097.

Takeo M, Lee W, Ito M. Wound Healing and Skin Regeneration. Cold Spring Harb Perspect Med 2015; 5: 1-12.systematic review. Burns, v. 40, n. 2, p. 177–190, mar. 2014. doi: 10.1101/cshperspect.a023267

Vloemans AF, et al. Optimal treatment of partial thickness burns in children: a systematic review. Burns. 2014 Mar;40(2):177–90. doi: 10.1016/j.burns.2013.09.016. Epub 2013 Nov 26. PMID: 24290852.

Whitaker IS, et al. The use of Biobrane® by burn units in the United Kingdom: A national study. Burns, v. 33, n. 8, p. 1015–1020, dez. 2007. doi: 10.1016/j.burns.2006.11.017. doi: 10.1016/j.burns.2006.11.017

World Health Organization (2018). Burns. https://www.who.int/news-room/fact-sheets/detail/burns.

Yarboro DDA, Comparative Study of the Dressings Silver Sulfadiazine and Aquacel Ag in the Management of Superficial Partial-Thickness Burns. Advances in Skin & Wound Care, v. 26, n.6, p. 259–262, jun. 2013. doi: 10.1097/01.ASW.0000431084.85141.d1

Yoshino Y. et al. Wound/Burn Guidelines Committee. The wound/burn guidelines - 6: Guidelines for the management of burns. J Dermatol. 2016 Sep;43(9):989–1010. doi: 10.1111/1346-8138.13288. doi: 10.1111/1346-8138.13288

